# Complex pathways to ceftolozane-tazobactam resistance in clinical *Pseudomonas aeruginosa* isolates: a genomic epidemiology study

**DOI:** 10.1101/2025.05.13.25327501

**Authors:** Hoai-An Nguyen, Anton Y. Peleg, Jiangning Song, Jessica A. Wisniewski, Luke V. Blakeway, Gnei Z. Badoordeen, Ravali Theegala, Nhu Quynh Doan, Matthew H. Parker, David L. Dowe, Nenad Macesic

**Author notes:** Corresponding Author: Nenad Macesic, Department of Infectious Diseases, The Alfred Hospital and School of Translational Medicine, Monash University, Level 1, The Alfred Lane House, 11 Alfred Ln, Melbourne, VIC 3004, Australia. No longer at the institution where the work was performed (Jessica A. Wisniewski, Luke V. Blakeway, Gnei Z. Badoordeen, Ravali Theegala).

## Abstract

**Objectives:** We aimed to conduct a comprehensive genomic analysis of ceftolozane/tazobactam (C/T) resistance mechanisms in *Pseudomonas aeruginosa* by combining novel institutional data with publicly available sequencing data.

**Methods:** We analysed 1,682 *P. aeruginosa* isolates, comprising 339 isolates from Alfred Hospital (Melbourne, Australia) and 1,343 isolates from six public datasets. All isolates underwent whole-genome sequencing and C/T broth microdilution (BMD) susceptibility testing. We assessed previously reported intrinsic and acquired resistance mechanisms. We then conducted a genome-wide association study (GWAS) and machine learning analysis to identify novel genes associated with resistance. We then evaluated the impact of mutations in these genes on MIC values and ceftolozane binding affinity.

**Results:** Among 1,682 *P. aeruginosa* isolates representing 527 distinct sequence types, 343/1,682 (20.4%) were C/T-resistant. Carbapenemase genes were detected in 206/1,682 (12.2%) isolates. Mutations in previously reported resistance-associated genes (*ftsI, mpl, ampD, ampC, ampR, oprD)* were more frequent in resistant isolates but were also found in almost all susceptible isolates. Successive mutations conferred additive increases in MIC. Combined GWAS and machine learning analyses *a priori* identified five key genes significantly associated with resistance: *ftsI*, *ampR*, *ampC*, PA3329, and PA4311. Molecular docking simulation revealed that the R504C mutation in penicillin-binding protein 3 (PBP3), which is encoded by *ftsI*, reduced binding contacts and hydrogen bonds with ceftolozane, significantly decreasing binding affinity (*P*=0.016).

**Conclusions:** Our analysis of 1,682 *P. aeruginosa* genomes demonstrated complex pathways to C/T resistance and showed that *ftsI* may play an underappreciated role. We discovered two previously unidentified genes associated with C/T resistance, whose function remains to be determined.

## Introduction

*Pseudomonas aeruginosa* is a key Gram-negative pathogen, accounting for 9.4% of global antimicrobial resistance (AMR)-attributable deaths in 2021^1^. Multidrug-resistant (MDR) *P. aeruginosa* forms 15-30% of total *P. aeruginosa* infections^2^, severely limiting available treatment options. Consequently, ceftolozane-tazobactam (C/T) has emerged as a first-line treatment for MDR *P. aeruginosa* infections^3^. However, rates of C/T resistance in MDR *P. aeruginosa* can be as high as 18.4%^4^. This highlights the urgent need to understand C/T resistance mechanisms, in order to rapidly identify resistant isolates and better control their spread.

There are multiple pathways contributing to C/T resistance^5^. Of these, structural alterations in AmpC β-lactamase represent most common mechanism^6–8^. AmpC hyperproduction, resulting from mutations in regulatory genes (*ampD*, *ampR*, *dacB,* and *mpl*), also confers resistance^6,7^. Acquired AMR genes such as carbapenemases or extended-spectrum β-lactamases represent another major mechanism due to their ability to hydrolyse ceftolozane^8,9^.

Previous studies on C/T resistance have largely focused on case series and well-described mechanisms^7,8,10^. However, the complex interplay between different resistance mechanisms remains poorly understood, and the identification of novel resistance determinants has been limited. To address these gaps, we aimed to comprehensively determine C/T resistance mechanisms using novel data from our institution combined with all publicly-available *P. aeruginosa* genomes with matched C/T susceptibility phenotypes. Specifically, we aimed to assess prevalence of known resistance mechanisms, then identify novel mechanisms through a genome-wide association study (GWAS) and machine learning (ML) approaches (**Figure 1**).

**Figure 1.**
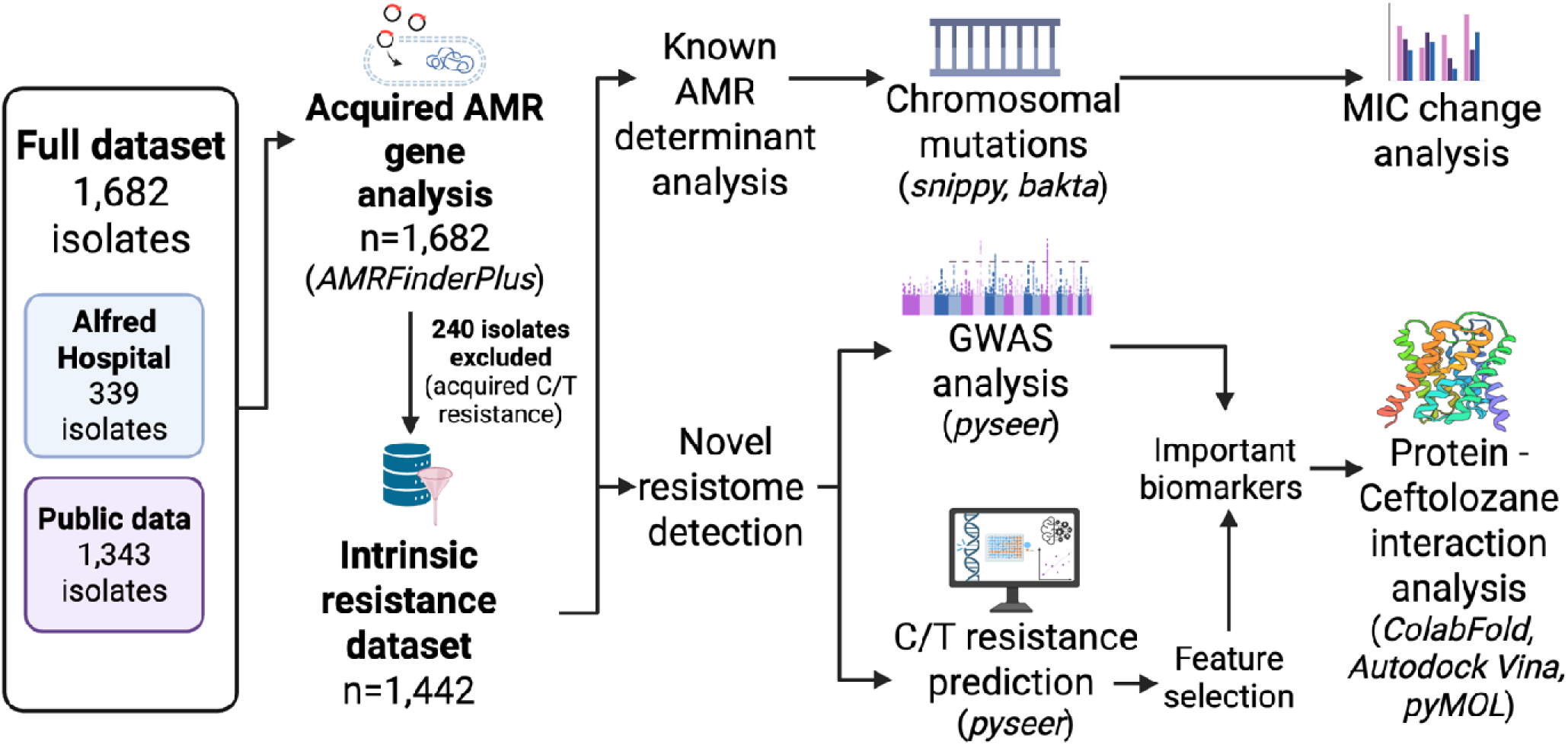
Overall workflow of the study. This figure was created with BioRender.com.

## Materials and methods

### Bacterial isolate collection

The study was reviewed and approved by the Alfred Hospital Ethics Committee (Project No. 185/21). We analysed 339 *P. aeruginosa* isolates collected between 2004-2021 at the Alfred Hospital (Melbourne, Australia), a 638-bed quaternary hospital with a cystic fibrosis and lung transplant state referral service. Consecutive *P. aeruginosa* isolates were recovered from clinical specimens. The isolates consisted of both matt and mucoid morphotypes and were stored in glycerol broth at –80°C. Repeat isolates from the same patient were included if they had a different morphotype and/or susceptibility profile (as determined by Vitek 2; BioMérieux, France) and were collected more than a week apart. All isolates underwent short-read sequencing (Illumina) (see **Supplementary Methods**).

We identified six public datasets containing both whole-genome sequencing (WGS) data and antimicrobial susceptibility testing (AST) results^5,11–15^. To ensure standardised resistance determination, we included only datasets where minimum inhibitory concentrations (MIC) were determined using broth microdilution (BMD), the reference method. The in-house sequence files have been deposited in NCBI project ID PRJNA1220180.

### Antimicrobial susceptibility testing

For our in-house isolates, we conducted BMD using the ThermoFisher Sensititre plate (code: DKMGN) to determine C/T MIC values. We used the European Committee on Antimicrobial Susceptibility Testing (EUCAST) v15.0 breakpoints (MIC ≤ 4mg/L) to classify isolates as ‘susceptible’ or ‘resistant’^16^.

### WGS data processing

*De novo* short-read assembly was generated using Unicycler v0.5.0^17^. Species confirmation and multi-locus sequence typing (MLST) were conducted via Pathogenwatch (https://pathogen.watch). Acquired AMR genes were detected using AMRFinderPlus v3.11.18^18^. For chromosomal mutations analysis, we assessed mutations in ten genes previously linked to C/T resistance, as described by Cortes-Lara et al.^5^. Additionally, we included *oprD* due to its involvement in resistance following C/T exposure (see Supplementary Methods)^7^.

### Identifying variants associated with C/T resistance

We conducted GWAS on susceptible/resistant phenotypes using a mixed-effects model implemented in pyseer v1.3.10^19^. Unitigs were used as testing variants, with a phylogeny-based pairwise distance matrix incorporated to control for population structure (see Supplementary Methods). Association mapping was performed using the annotated PAO1 reference genome (Genbank: GCA_000006765.1). To reduce false positives and computational load, we applied a two-step filtering strategy: first identifying GWAS-significant variants, then retaining only those that also contributed to ML-based C/T resistance prediction (see Supplementary Methods).

### Binding affinity analysis

We assessed the binding affinity between ceftolozane and proteins encoded by GWAS-significant genes, comparing both wild-type and mutant variants. 3D structures of proteins were predicted using ColabFold, an AlphaFold-based tool^20^. Molecular docking simulations were performed using AutoDock Vina v1.2.5^21^, and results were visualised using PyMOL^22^.

## Results

### Data characteristics

We analysed 1,682 *P. aeruginosa* genomes from four continents (Asia, Europe, North America, Oceania) (**Supplementary File 1**), including Alfred Hospital isolates (339/1,682, 20.2%) and six public datasets (1,343/1,682, 79.8%)^5,11–15^. C/T resistance was noted in 343/1,682 (20.4%) isolates. We detected 527 sequence types (STs) (**Supplementary Figure 1**). *P. aeruginosa* ST235, a well-recognised global MDR lineage was most frequent (163/1,682, 9.7%) and accounted for 78/343 (22.7%) of C/T resistant isolates. Resistance rates in common ST groups can be found in **Supplementary Table 1**.

### Prevalence of acquired AMR genes

We first analysed acquired C/T resistance determinants. Carbapenemase genes were noted in 206/1,682 (12.2%) isolates (**Supplementary Table 2**), including serine carbapenemase (*bla*_GES_, *bla*_KPC_) and metallo-β-lactamase genes (*bla*_IMP_, *bla*_VIM_, *bla*_NDM_). Carbapenemase acquisition strongly predicted C/T resistance, with 202/206 (98.1%) carbapenemase-positive isolates being C/T resistant. Only *bla*_GES-5_ was found in C/T-susceptible isolates (4/206, 1.9%). The most common carbapenemase was *bla*_NDM-1_ (37/206, 18.0%), followed by *bla*_IMP-4_ (31/206, 15.0%), *bla*_IMP-1_ (29/206, 14.1%), *bla*_VIM-2_ (25/206, 12.1%) and *bla*_GES-5_ (22/206, 10.7%). Dual carbapenemases were identified in six isolates. The *bla*_OXA-50_ family was excluded from analysis as these enzymes are intrinsic to *P. aeruginosa*^23^. Among acquired *bla*_OXA_ genes, *bla*_OXA-10_, *bla*_OXA-14_, *bla*_OXA-141_, *bla*_OXA-210_, and *bla*_OXA-796_ were significantly more prevalent in C/T-resistant isolates (**Supplementary Figure 2**). VEB-type β-lactamases were detected only in C/T-resistant isolates (8/343, 2.0%), with 4/8 co-harboring additional carbapenemases: two with *bla*_KPC-2_, one with *bla*_VIM-5_, and one with *bla*_NDM-1_.

### Analyses of known chromosomal C/T resistance mechanisms

We then assessed prevalence of mutations in a curated list of eleven intrinsic genes associated with C/T resistance (*ampC* and its regulators, mexAB-oprM overexpression pathway, *ftsI, galU* and *oprD*, **Figure 2A-C, Supplementary Table 3**)^5^. We specifically excluded isolates with acquired resistance determinants that likely conferred C/T resistance alone (carbapenemase genes, VEB β-lactamases, acquired OXA β-lactamases with significantly higher prevalence in resistant isolates), as the impact of additional mutations would be unclear. This resulted in a subset of 1,447 isolates (including 115 resistant isolates), which we will refer to as the ‘intrinsic resistance’ dataset. Mutations in genes associated with resistance were noted in all resistant isolates but also in 1,323/1,332 (99.3%) susceptible isolates, however the majority of these have previously been described as natural polymorphisms (**Supplementary Table 3**)^5^.

**Figure 2.**
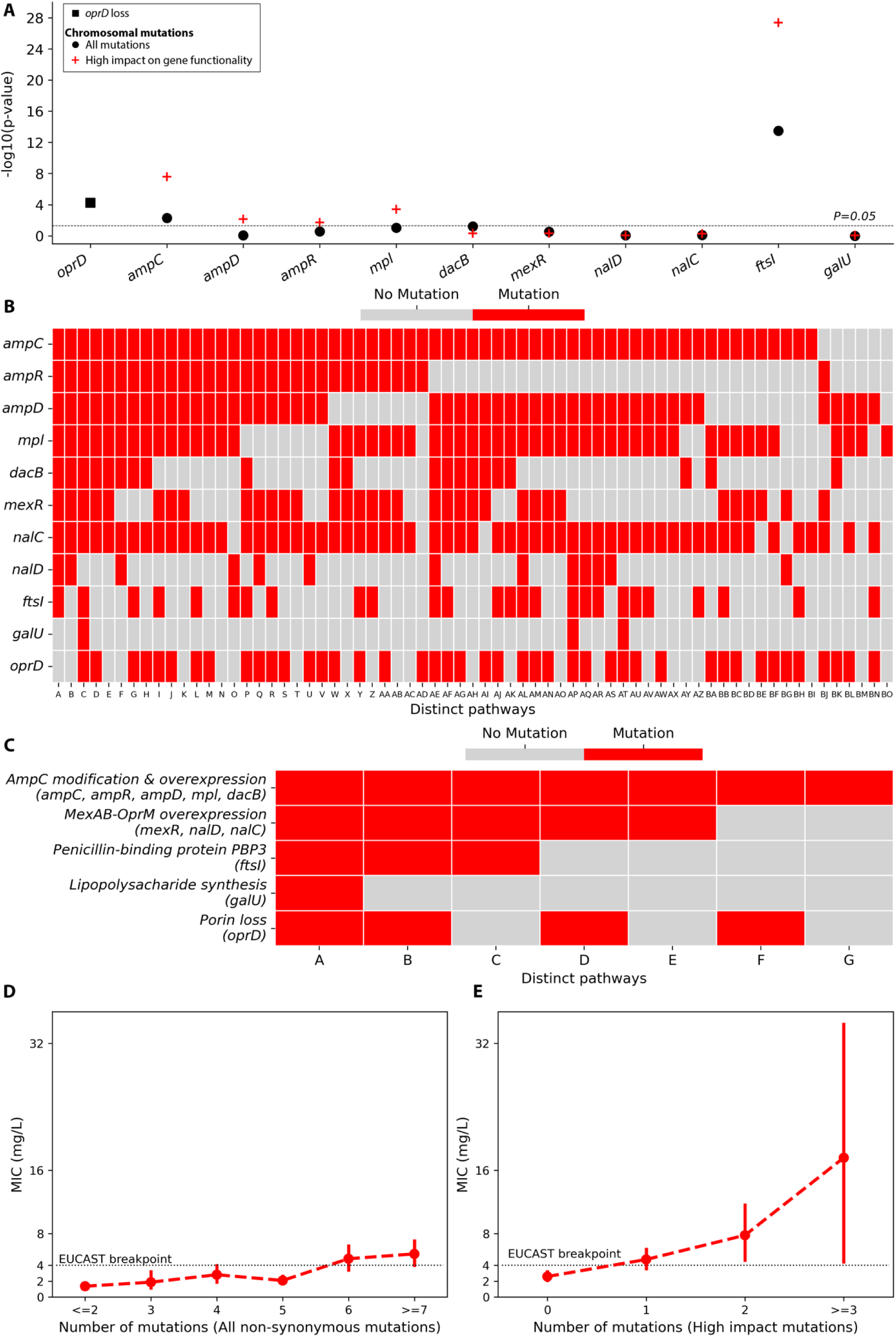
**(A) Impact of mutations in chromosomal genes known to affect C/T susceptibility.** P-values were calculated using the Chi-square test. **(B) Pathways to C/T resistance at gene level.** Each unique combination of mutations in genes associated with C/T resistance is shown as a unique pathway. **(C) Distinct pathways to C/T resistance, collapsed to resistance mechanisms.** Here, the genes associated with C/T resistance are collapsed into mechanistic groupings. Each unique combination of mutations in mechanistic groupings is shown as a unique pathway. **(D, E) MIC values with respect to the cumulation of mutations (all non-synonymous mutations [D] vs high-impact mutations only [E]) in chromosomal genes known for C/T resistance.** Dots and vertical bars indicate mean and standard deviation (SD) of MIC values, respectively.

We therefore focused on mutations with a previously reported high impact on C/T susceptibility. These were significantly more frequent in resistant than susceptible isolates (in order of significance): *ftsI* (24/115 resistant vs. 22/1,332 susceptible isolates, *P*=4.11×10^−28^), *ampC* (19/115 vs 55/1,332, *P*=2.58×10^−8^), *mpl* (12/115 vs. 44/1,332, *P*=3.82×10^−4^), *ampD* (9/115 vs 37/1,332, *P*=0.007), and *ampR* (13/115 vs 73/1,332, *P*=0.02). In terms of specific mutations, for *ftsI* the R504C mutation was most prevalent (21/115 [18.3%] resistant vs 17/1332 [1.3%] susceptible isolates). Notably, it was distributed across 18 unique STs in the resistant group. The previously reported F533L and E466K mutations were not found in C/T-resistant isolates^24,25^. For known *ampC* mutations^26^, E247G was detected in one resistant and one susceptible isolate. Deletions in the G229-E247 region (△G at L237, △P241-A246, and △R238-L244) were found in four resistant isolates and no susceptible isolates.

Inactivated *oprD* was also more frequent in resistant isolates (77/115 [66.9%] vs 626/1,332 [47,0%], *P*=6.03X 10^−5^). Frameshift due to indels was the most common mutation in both resistant (26/115, 22.6%) and susceptible (249/1,332, 18.7%) isolates. For other canonical C/T resistance genes, mutations in the *mexAB-oprM* efflux pump regulators (*nalC*, *nalD*, and *mexR*) were very frequent in the resistant group, occurring in 111/115 (96.5%). Mutations in *galU* and *dacB* were noted in 3/115 (2.6%) and 39/115 (33.9%) resistant isolates, respectively.

Analysis of combinations of mutations in C/T resistance-associated genes revealed 67 distinct mutational pathways in resistant isolates (**Figure 2B**). Multiple mutations were common, with only 1/67 (1.5%) pathways involving a single mutation. Notably, all resistant isolates harboured at least one mutation in either *ampC* or its overexpression regulators (*ampR*, *ampD*, *mpl, dacB*). At the AMR mechanism level, collapsing *ampC*-related genes and *mexAB-oprM*-related genes into functional groups revealed seven distinct resistance pathways (Figure 2C). Among these, only modifications in AmpC and its regulatory genes emerged as a standalone pathway capable of conferring C/T resistance. While AmpC overexpression represents the main mechanism of resistance to C/T, additional mechanisms are often required to reach high-level resistance, highlighting the complexity of the mutational resistome in *P. aeruginosa*.

We next investigated the impact of multiple mutations on MIC, firstly by including both low- and high-impact mutations (**Figure 2D**, **Supplementary Figure 3A**), then high-impact mutations only (**Figure 2E**, **Supplementary Figure 3B**). In both analyses, we observed an increasing trend in MIC with a higher number of mutations. High-impact mutations were associated with higher MIC increases, with a single mutation able to achieve resistant MICs (mean: 4.73 mg/L, SD: 11.64, **Figure 2E**) per EUCAST breakpoints (4 mg/L). When considering both low- and high-impact mutations, six or more mutations were required to achieve the same effect (**Figure 2D)**.

### Identifying novel C/T resistance mechanisms through GWAS and ML analysis

Using the ‘intrinsic resistance’ dataset (n=1,447), we conducted GWAS and identified 311 significant variants out of 2,698,635 tested unitigs (**Supplementary Figure 4**). Mapping to the PAO1 reference genome annotated 280 unitigs corresponding to 73 genes (**Supplementary Table 4**). Top associated genes included *ftsI* (*P*=5.05×10^−23^), *hsbR* (*P*=5.85×10^−20^), *ampR* (*P*=3.52×10^−18^), *ampC* (*P*=6.50×10^−15^), and several hypothetical proteins, including PA3329 (*P*=1.47×10^−15^), PA4311 (*P*=6.86×10^−15^), and PA2418 (*P*=1.58×10^−13^) (**Figure 3A**).

**Figure 3.**
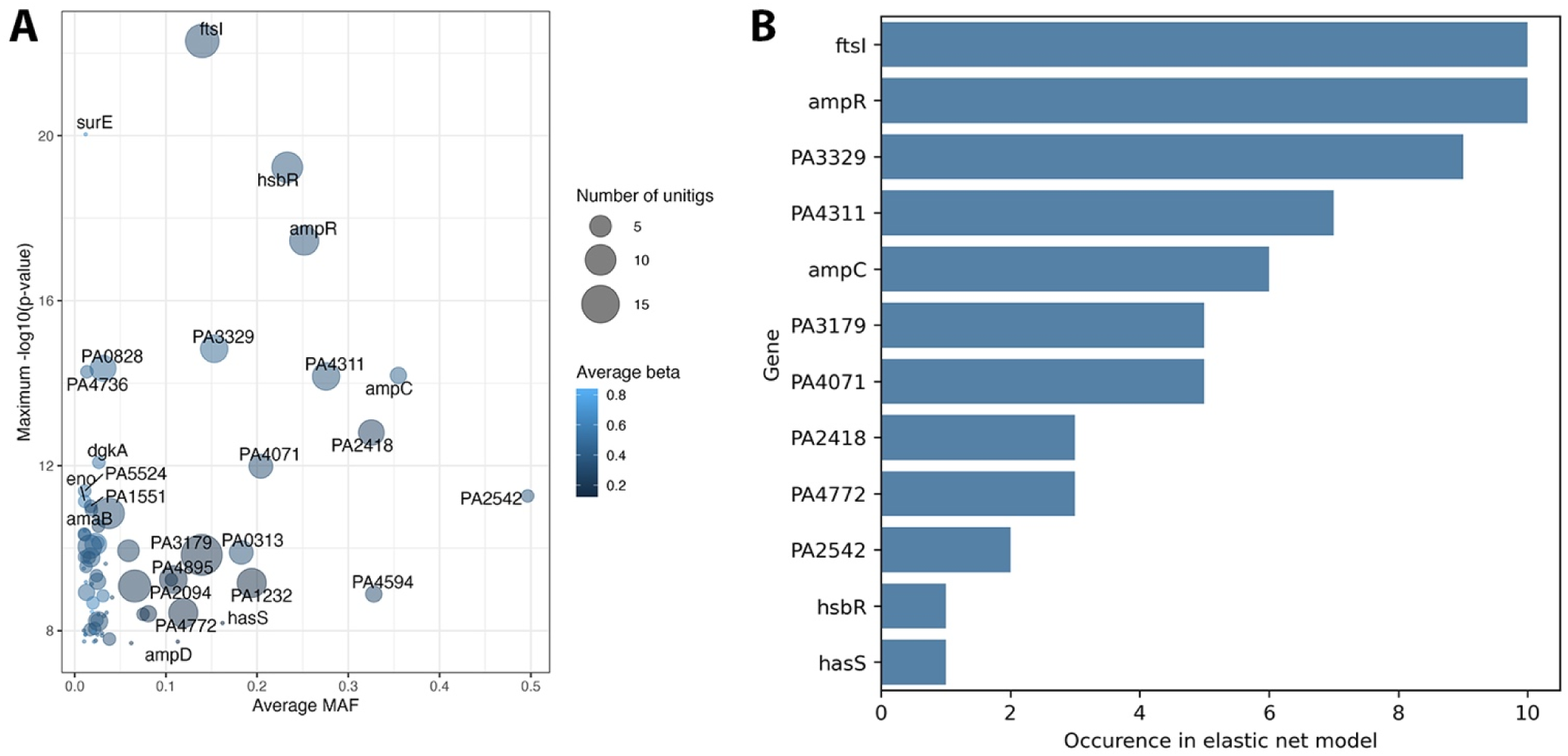
**(A) Genes selected by GWAS study, with the maximum P-value (y-axis) and the average minimum allele frequency (MAF) (x-axis). (B) Genes (y-axis) selected by both GWAS and ML approach, and their occurrence in ML models (x-axis).** Only genes with MAF > 0.1 are shown.

We then developed ML models to predict binary C/T resistance using the same dataset, achieving an average area under receiver operator curve (AUROC) of 0.65, and log-loss of 0.26 (**Supplementary Figure 5**). Across all model iterations, 386 unitigs were selected. To refine the list of candidates, we intersected GWAS-significant unitigs with those selected by the models, applying thresholds of a minimum allele frequency above 0.1 and presence in more than 5 model iterations (**Figure 3B**, **Supplementary Table 4**). This filtering yielded five key genes: *ftsI* and *ampR* were selected in all models, while PA3329, PA4311, and *ampC* appeared in 9/10, 7/10, and 6/10 models, respectively.

### Binding affinity assessment for significant variants

*ftsI* emerged as a key determinant of C/T resistance in analysis of known resistance mechanisms, as well as GWAS/ML analysis. These findings and the fact that PBP3 is a target of ceftolozane prompted investigation of mutation impacts on *in silico* binding affinity^27^. We identified four non-synonymous mutations in GWAS-significant unitigs mapped to *ftsI*: R504C (n=38), F507I (n=1), T514A (n=1), and R504H (n=1). Among these, only R504C was significantly associated with resistant isolates (21/115 [18.3%], *P*=2.30×10^−26^) while the others were exclusively found in susceptible isolates. Molecular docking analysis revealed significantly higher binding affinity between ceftolozane and wild type PBP3 compared to mutant PBP3 (R504C) (-3.520 vs -2.814 kcal/mol, *P*=0.016) (**Supplementary Figure 6**), with loss of contact at A244 and single hydrogen bonds at Q265 and T267 (cf. double bonds in wild type PBP3) (**Figure 4**). These changes potentially explain reduced drug binding and consequent resistance.

**Figure 4.**
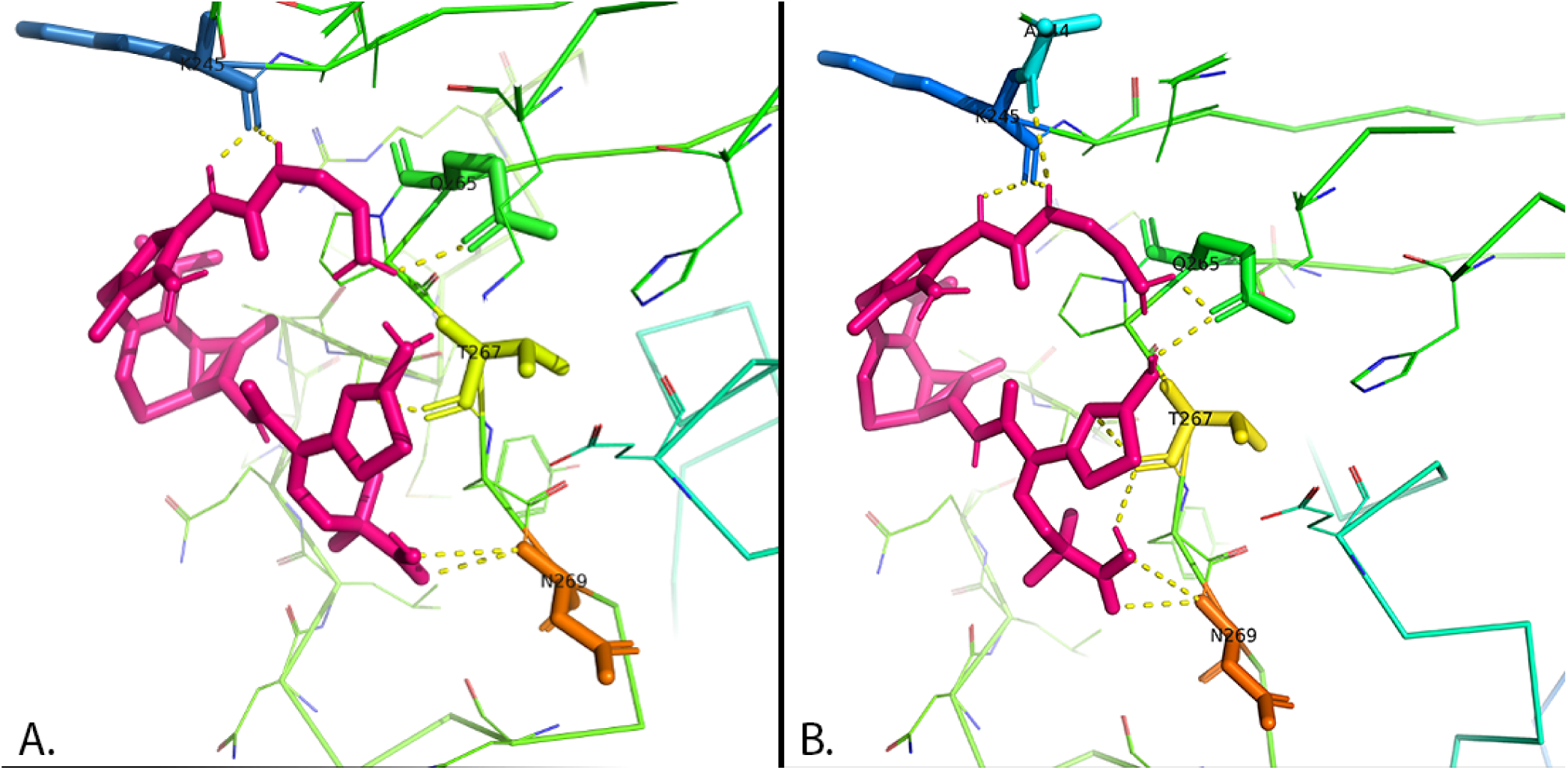
**3D structure visualization of the interaction of ceftolozane (pink) to R504C mutant PBP3 (Panel A) and wild type PBP3 (Panel B).** Dashed yellow lines indicate hydrogen bonds. The R504C mutant (**Panel A**) shows loss of contact at A244 and single hydrogen bonds at positions Q265 and T267 (cf. double bonds in wild type PBP3 – **Panel B**).

We extended our docking analysis to the novel proteins encoded by PA3329 and PA4311. The PA3329-encoded protein showed no meaningful binding affinity to ceftolozane (mean: 924.393 kcal/mol, SD: 288.148). The PA4311-encoded protein demonstrated binding affinity (-1.493 kcal/mol, 1.459) with potential binding sites at D116, Q271, R292 and G294 (**Supplementary Figure 7**).

### Putative functions of novel C/T resistance determinants

To understand putative functions of PA3329 and P4311, we examined their genomic neighbourhood, predicted Gene Ontology functions^28^ and conducted a database and literature search (see **Supplementary Methods** and **Supplementary Table 5)**. PA3329 is located in an eight-gene operon (PA3327-PA3334) that includes *fabH2* (beta-keto-acyl-ACP synthase) and *acp3* (acyl carrier protein), suggestive of lipid/lipopolysaccharide synthesis^29,30^ (**Supplementary Figure 8)**. The predicted Gene Ontology terms for PA3329 indicate a role in cellular anatomical structure **(Supplementary Table 5)**. PA3329 has also been implicated in the CreBC two-component system which is involved in β-lactam stress response^31^. PA4311 is annotated as a glycosyltransferase^32^ (https://www.pseudomonas.com/feature/show?id=111460). The predicted Gene Ontology terms also indicate glycosyltransferase activity and a role in cellular anatomical structure (**Supplementary Table 5**). In *P. aeruginosa*, glycosyltransferase reactions are essential for lipopolysaccharide synthesis, which is located in the outer leaflet of the outer membrane^33^. Based on this evidence, we hypothesise that both PA3329 and PA4311 are involved in outer membrane remodelling that affects ceftolozane influx.

## Discussion

This comprehensive genomic analysis of 1,682 *P. aeruginosa* isolates has revealed multiple mechanisms contributing to C/T resistance, including previously known resistance determinants (*ftsI*, *ampC*, and *ampR*) and novel candidates (PA3329 and PA4311). *ftsI* emerged as a key contributor to C/T resistance across multiple analyses. These findings were underscored by several methodological strengths. Our study leveraged a large, geographically diverse dataset incorporating both novel and public genome sequences to provide robust statistical power and broad representativeness. Additionally, our multi-modal approach, which combines GWAS, ML, and molecular docking, enabled cross-validation of findings through complementary methodologies.

Ceftolozane is a potent inhibitor of PBP3 encoded by *ftsI*^27^. Our study noted that mutations in *ftsI* were highly associated with C/T resistance, being present in 24/115 resistant vs 22/1,332 susceptible isolates (*P*=4.11×10^−28^), specifically the R504C mutation. This was then further confirmed in our GWAS and ML analyses, which both identified *ftsI* as the top-ranking contributor to C/T resistance. Our docking analysis then demonstrated that mutant *ftsI* (R504C) significantly reduced ceftolozane binding affinity, providing a mechanistic explanation for resistance. The role of PBP3 in β-lactam resistance has been increasingly recognised, not only in *P. aeruginosa*^7^, but also across various Enterobacterales^34,35^. Although the R504C mutation was previously documented to emerge following C/T exposure^10^, its importance in C/T resistance has potentially been underappreciated, as research has primarily focused on β-lactamases^8,36^. Furthermore, its co-occurrence with other mutations may have obscured its individual contribution to resistance^7,8,10^, highlighting the value of GWAS approach in identifying specific resistance determinants.

Our analysis of genes previously associated with C/T resistance^5^ showed that they likely have differing contributions when studied in a diverse international dataset. We applied a multi-modal approach to assess the contribution of resistance genes by applying both a traditional genomic analysis looking at genes associated with C/T resistance in the literature, and GWAS and ML approaches to allow us to identify key resistance genes *a priori.* Mutations in *ampC* and its regulator genes were found in all resistant isolates. Amongst these some were confirmed to have strong contributory effects (e.g. *ampC* and *ampR*) but others contributed less (e.g. GWAS suggested limited contributions from *ampD*). In some C/T resistance genes we detected very few mutations (e.g., *galU* in 2.6% of resistant isolates). These findings highlight the complex nature of C/T resistance mechanisms in *P. aeruginosa*. To try to account for some of these interactions, we assessed the impact of multiple mutations and noted stepwise increases in MIC with the number of mutations, both when considering all non-synonymous mutations and when considering high-impact mutations alone.

The contribution of horizontally acquired β-lactamase genes to C/T resistance has been well described^8,9^, specifically key carbapenemases including *bla_VIM_*, *bla_IMP_*, *bla_NDM_*, and *bla_KPC_*. While we noted these carbapenemases in 12.2% isolates in our study, we also showed that specific *bla*_OXA_ and *bla*_VEB_ genes may play an important role. Four *bla*_OXA_ genes (*bla*_OXA-14_, *bla*_OXA-141_, *bla*_OXA-210_, and *bla*_OXA-796_) and all *bla*_VEB_ genes were found exclusively in resistant isolates. These findings confirm prior studies that identified *bla*_OXA_ gene acquisition as a common cause of C/T resistance (particularly *bla*_OXA-14_)^8^, as well as noting an association between *bla*_VEB_ gene acquisition and C/T resistance^37^.

Our study had several limitations. Firstly, the retrospective nature of the analysis and potential sampling bias in the dataset may affect generalisability. Secondly, while docking analysis suggests reduced ceftolozane binding to mutant PBP3, in vitro studies are required to confirm these findings. Lastly, we hypothesised that PA3329 and PA4311 are involved in outer membrane remodelling however future functional validation is crucial to testing this hypothesis and determining their role in resistance. However, the emergence of mutations and interactions between multiple genes complicates the identification of key resistance drivers.

In conclusion, our study rigorously determined the genetic underpinnings of C/T resistance and revealed complex pathways leading to C/T resistance. *ampC* and its regulators were confirmed to play an important role but we demonstrated that *ftsI* may be a more significant contributor than previously recognised. We also identified two novel determinants whose functions remain to be determined. The hypotheses generated by our study present an opportunity for validation through both computational and experimental approaches to understand the functional mechanisms of these newly identified determinants.

## Data Availability

The in-house sequence files have been deposited in NCBI project ID PRJNA1220180

## Author contributions

HA.N, N.M. and A.Y.P. conceived the study. J.A.W. designed and supervised sampling and collection of bacterial isolates. J.A.W., L.V.B., G.Z.B, R.T., N.Q.D., and M.H.P. collected the bacterial isolates, performed bacterial characterization, conducted genome sequencing and broth microdilution. HA.N. preprocessed WGS data. HA.N., J.S., D.L.D., and N.M. conceptualised machine learning analyses. HA.N. conducted GWAS, developed machine learning models. HA.N. and N.M. analysed all results. HA.N. and N.M. wrote the manuscript with comments and feedback of all of the co-authors. All authors read and approved the manuscript.

## Acknowledgement

This work was supported by Monash eResearch Centre, including the M3 service. This work was also supported by use of the Nectar Research Cloud, a collaborative Australian research platform supported by the NCRIS-funded Australian Research Data Commons (ARDC).

