## Supplementary Materials for "Complex pathways to ceftolozane-tazobactam resistance in clinical *Pseudomonas aeruginosa* isolates: a genomic epidemiology study"

**Supplementary methods**

*Whole genome sequencing (WGS)*

The protocol for obtaining WGS data was previously described by Macesic et al.^1^. In brief, *P. aeruginosa* isolates were first cultured on cation-adjusted Mueller-Hinton II agar, then sub-cultured in cation-adjusted Mueller-Hinton broth (Becton-Dickinson). Genomic DNA was extracted using the GenFind V3 Reagent Kit (Beckman Coulter). Short-read sequencing libraries were prepared using the Nextera Flex DNA Library Prep Kit (Illumina), followed by 150 bp paired-end sequencing on the NovaSeq 6000 system (Illumina).

*Phylogenetic tree construction*

Pan-genome analysis was performed with Panaroo v1.2.4^2^, and a phylogenetic tree based on core gene alignment was constructed using IQ-TREE v2.0^3^. The resulting pairwise distance matrix was used to account for population structure in genome-wide association study (GWAS). The phylogenetic tree was visualized using the ggtree (v3.12.0) R package^4^.

*Chromosomal antimicrobial resistance (AMR) analyses*

We examined variants in 11 genes previously associated with ceftolozane/tazobactam (C/T) resistance^5^, including *ampC*, *ampR*, *ampD*, *mpl*, *dacB*, *mexR*, *nalC*, *nalD*, *ftsI*, *galU*, and *oprD*. We performed variant calling with Snippy v4.4.5 (https://github.com/tseemann/snippy), excluding all synonymous variants. Loss of OprD was assessed using PorinPredict v1.0.0^6^, and gene presence/absence was determined using Bakta v1.9.3^7^.

*Genome-wide association study*

We employed unitigs as the variant type for our GWAS analysis. Using unitig-caller (https://github.com/bacpop/unitig-caller) with the default k-mer length of 31, we generated a unitig database comprising 2,698,635 unitigs tested in the GWAS. Following pyseer’s approach for adjusting the significance threshold, we used the number of unique unitig patterns (2,311,870) to calculate the Bonferroni-corrected threshold, yielding *P*=2.16$\times$10^-8^.

*Predicting ceftolozane/tazobactam resistance with machine learning*

We utilised pyseer’s prediction module to develop models for C/T resistance prediction^8^. Specifically, we implemented Elastic Net models using unitig features, with the dataset split into training (80%) and testing (20%) sets, stratified by antimicrobial susceptibility testing (AST) labels. We ran the models ten times with different random splits, using the area under the receiver operating characteristic curve (AUROC) as the performance metric. Salient unitigs were defined as those with non-zero coefficients in the Elastic Net models. To refine our GWAS results, we identified the intersection between GWAS-significant variants and these salient unitigs.

*Protein structure prediction*

Wild-type protein sequences for the investigated genes were obtained from the Pseudomonas Genome Database (https://www.pseudomonas.com). Mutant sequences were generated by introducing specific amino acid substitutions. Protein structure prediction was performed using the ColabFold implementation of AlphaFold2 via their official Google Colab notebook (https://colab.research.google.com/github/sokrypton/ColabFold/blob/main/AlphaFold2.ipynb) with default parameters^9^.

*Assessment of putative functions of novel C/T resistance determinants*

For GWAS-significant genes with unknown functions, we first performed a database search on Pseudomonas.com^10^ to assess their genomic location and surrounding genes in the PAO1 genome^11^. Potential gene annotations were also retrieved from the database and cross-validated with predicted Gene Ontology (GO) functions. Here, we used the pretrained DeepGO-SE model^12^ to predict GO functions across three categories: molecular function, cellular component, and biological process. Protein sequences from the PAO1 reference genome served as model input. A probability threshold of 0.8 was applied to retain only high-confidence predictions. In addition, obsolete terms were excluded from the results. Lastly, we performed a literature search to identify any supporting evidence for the inferred functions of the candidate genes.

**Supplementary figures**

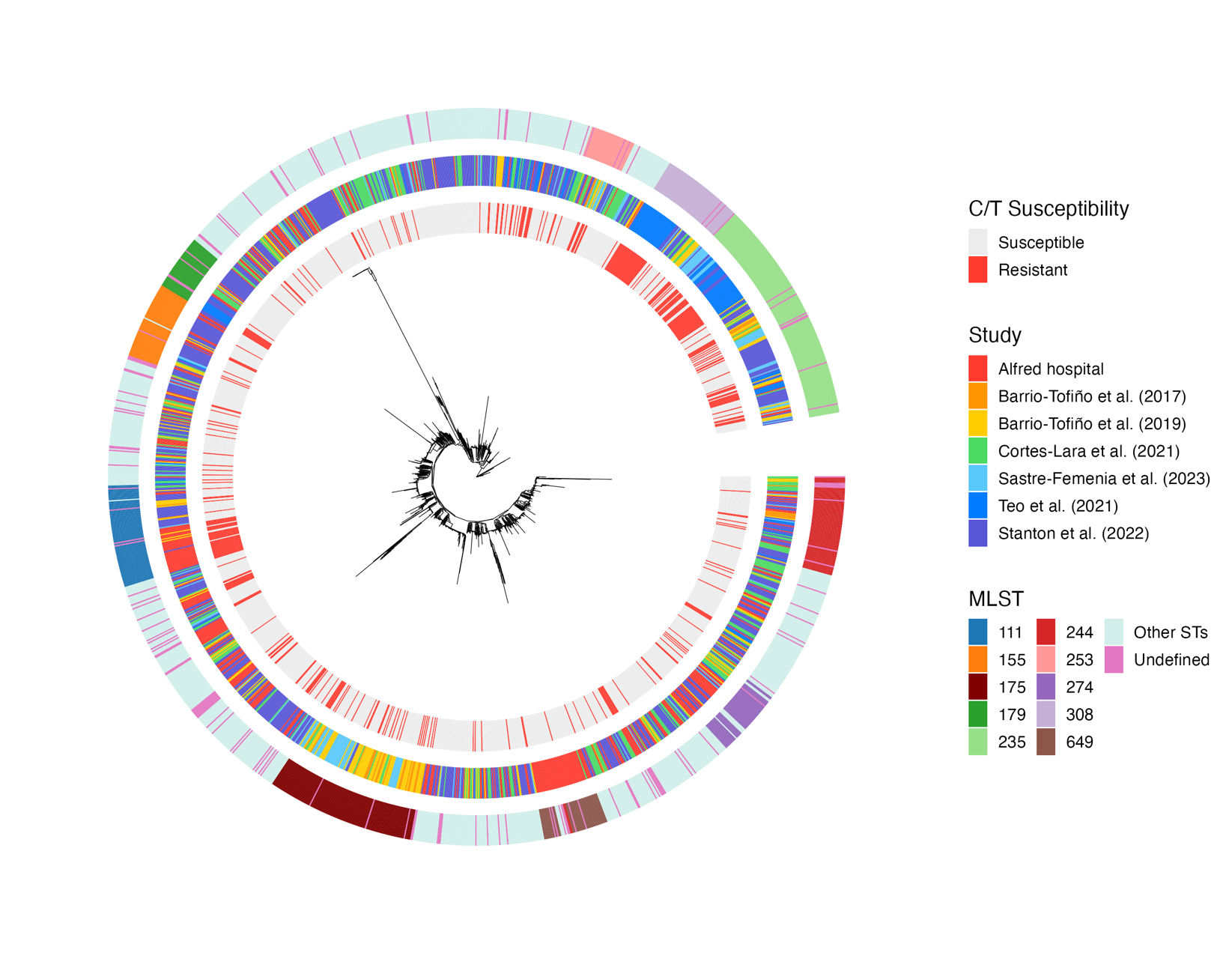

**Supplementary Figure 1. Phylogenetic tree of whole dataset.** From inside to outside, the rings depict ceftolozane/tazobactam (C/T) susceptibility, isolate dataset origin, and multi-locus sequence type (MLST) information, respectively.

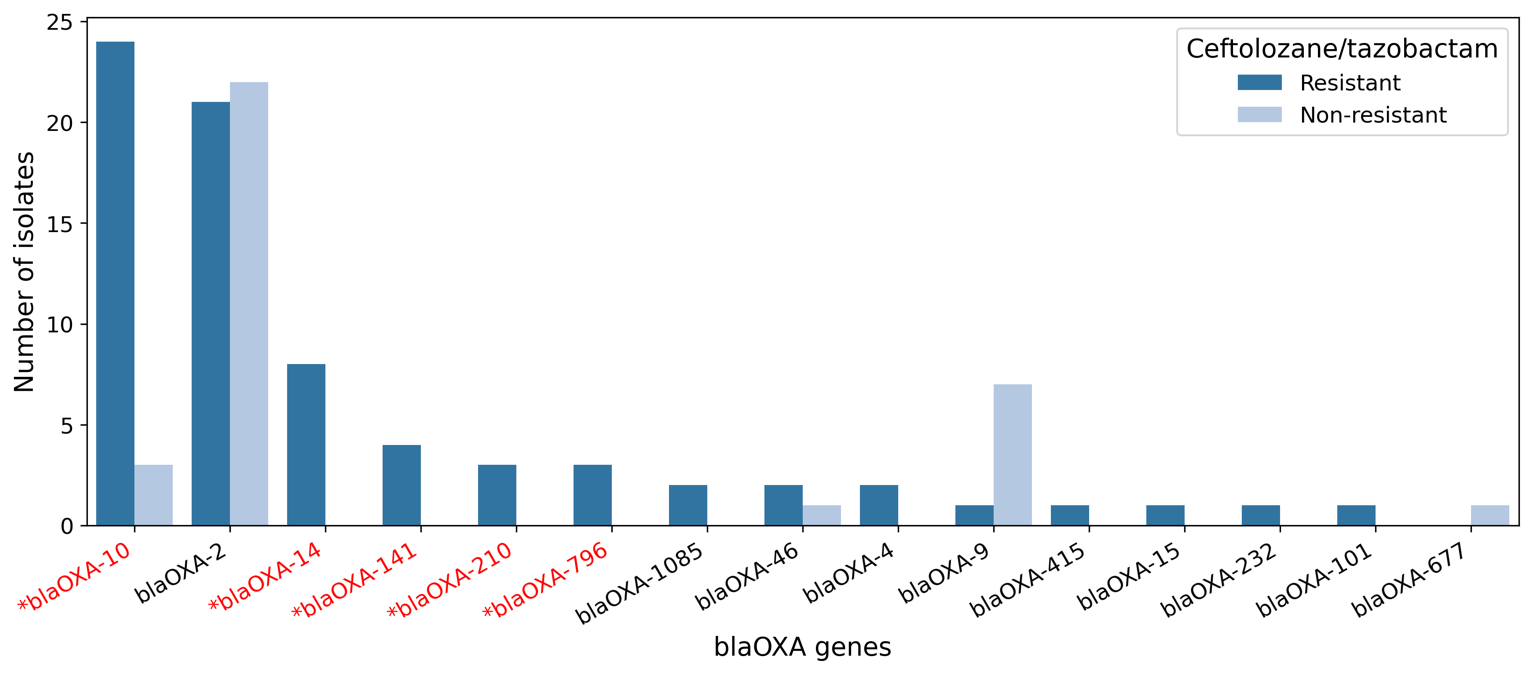

**Supplementary Figure 2. Occurrence of *bla*_OXA_ genes in the study population between ceftolozane-tazobactam susceptible and resistant isolates.** Red text with an asterisk (*) indicates *bla*_OXA_ genes significantly present in the resistant group (Chi-square test).

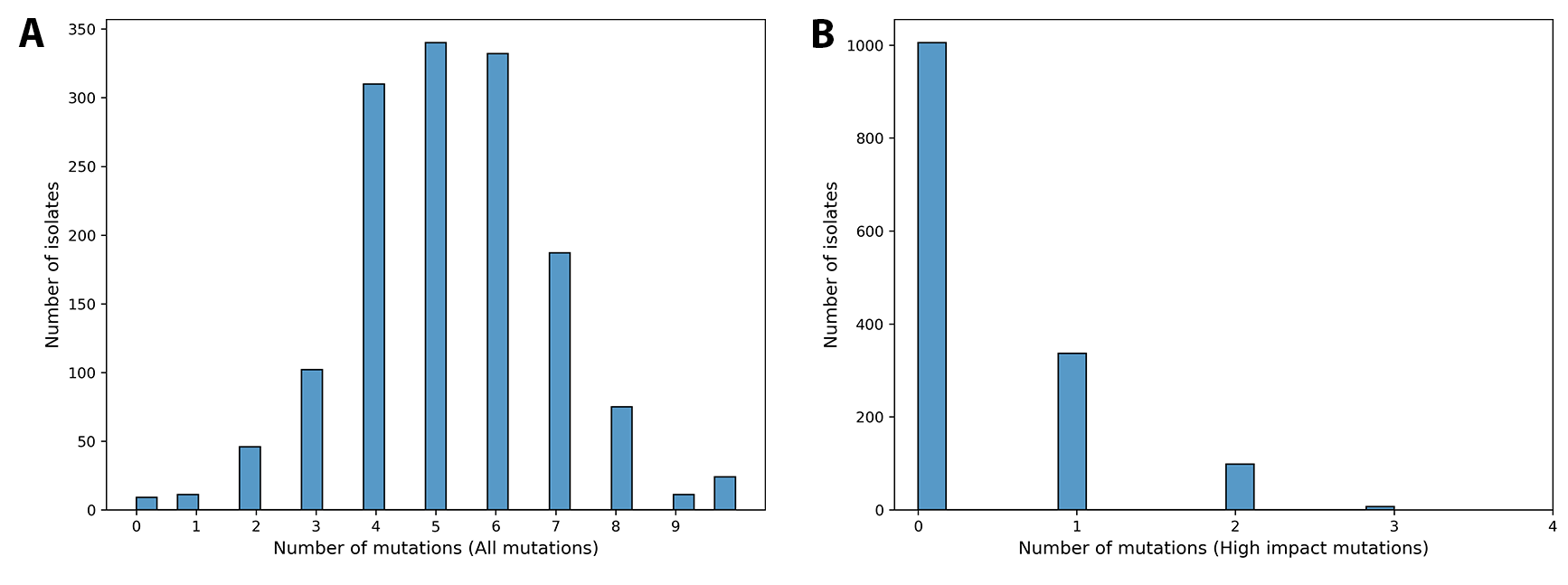

**Supplementary Figure 3. Total number of mutations carried by individual isolates of genes related to C/T resistance, considering all mutations (A) and only mutations having high impact on C/T resistance (B).**

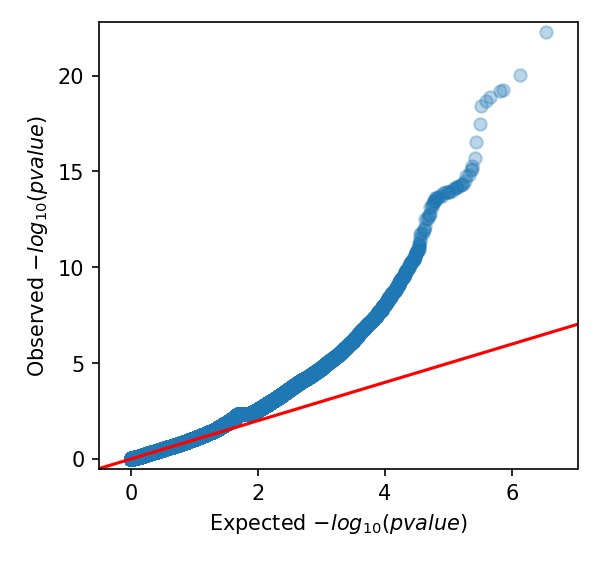

**Supplementary Figure 4. P-values of all tested variants in the GWAS study.**

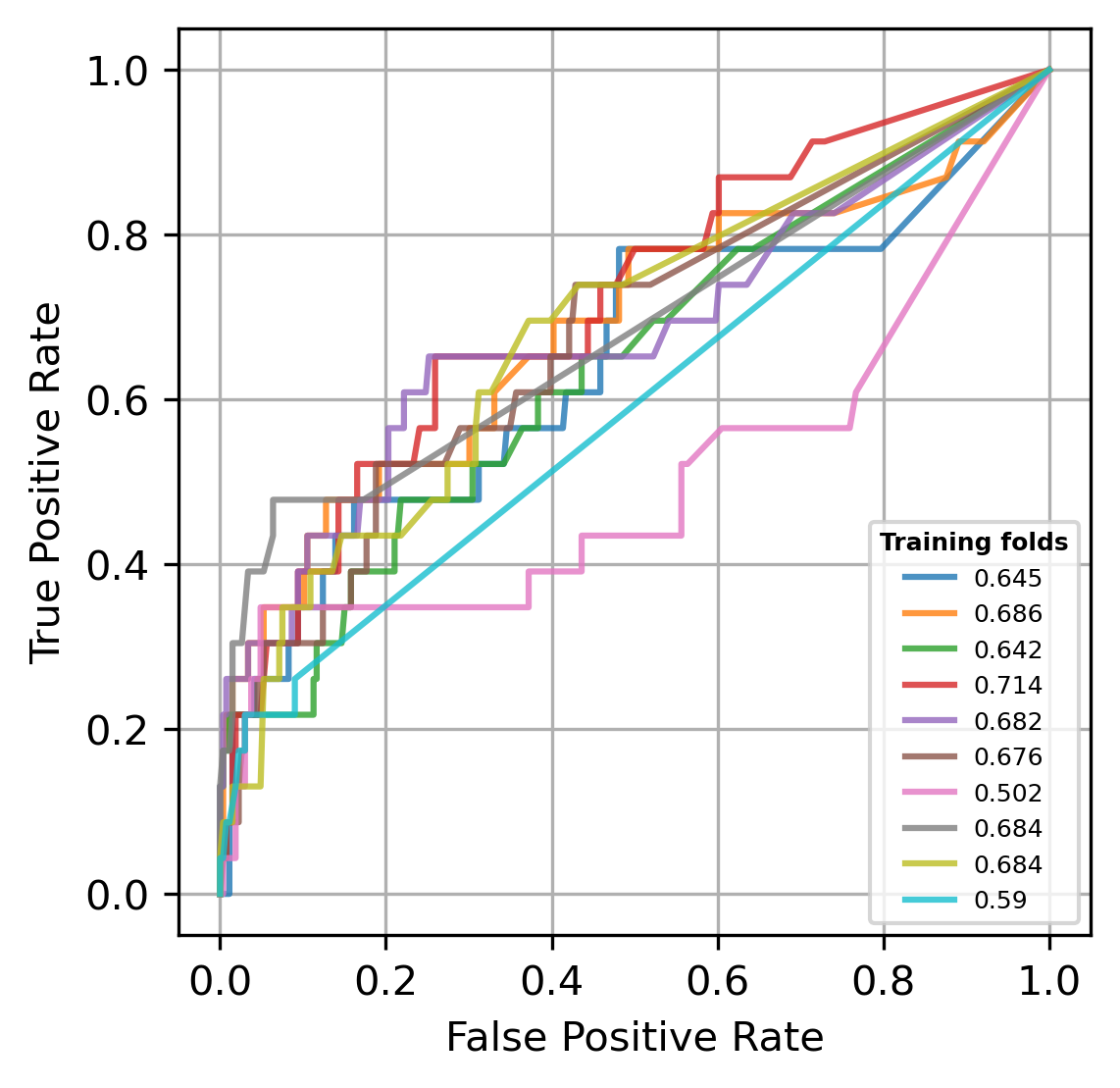

**Supplementary Figure 5. AUROC performance of models predicting C/T resistance across 10 runs.**

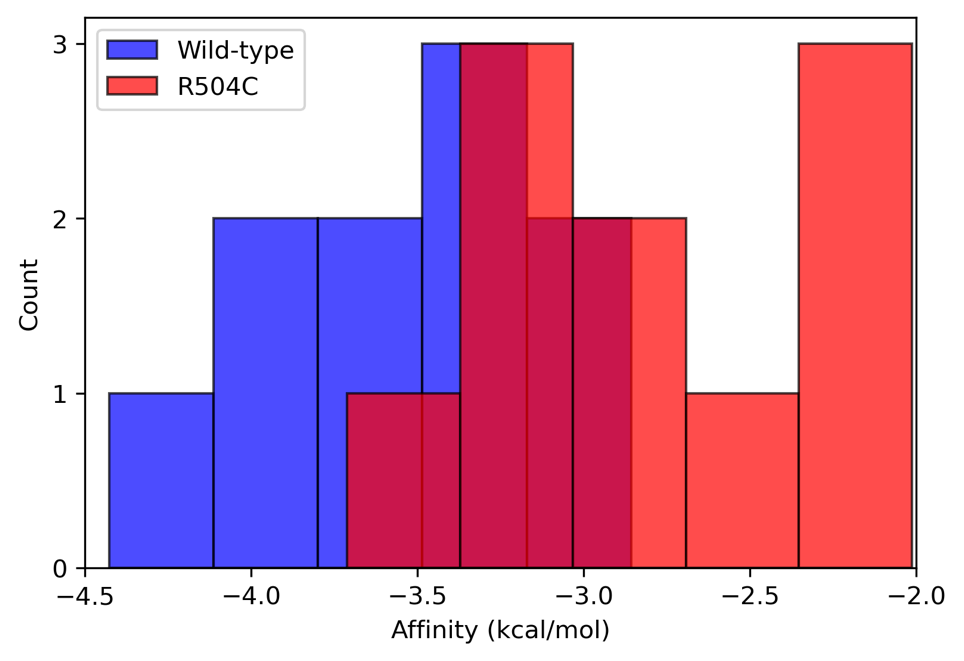

**Supplementary Figure 6. Affinity energy between the protein (Penicillin-binding protein 3 - PBP3) encoded by *ftsI* (blue: wild-type; red: R504C mutant) and ceftolozane across 10 random runs.**

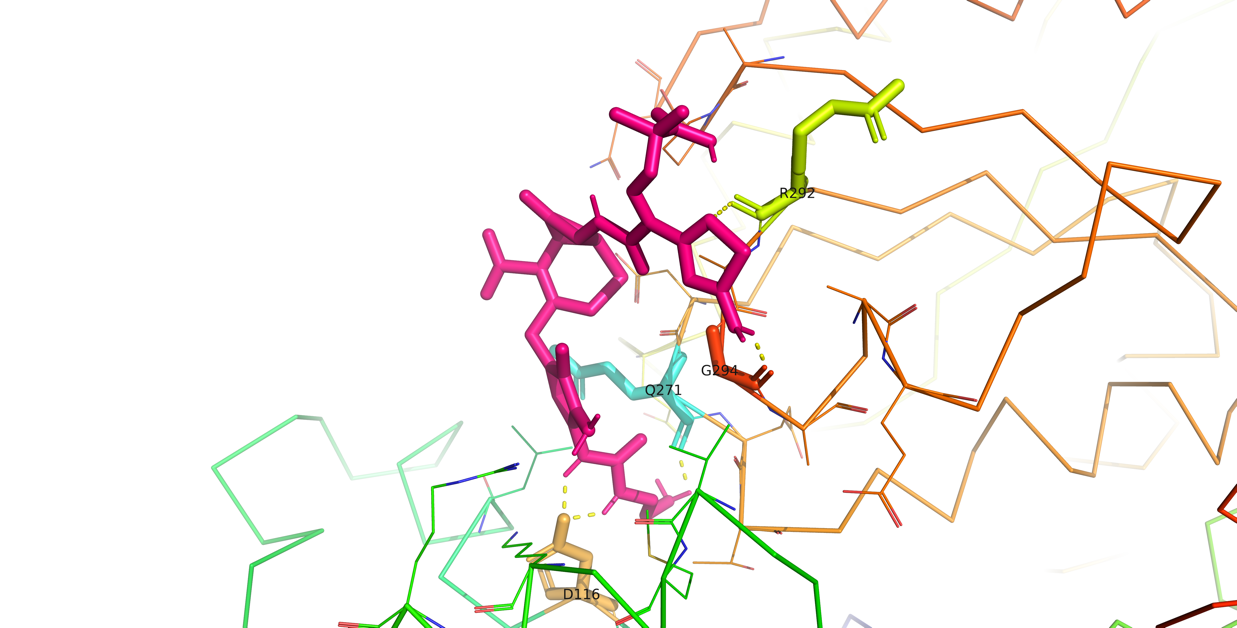

**Supplementary Figure 7. Binding interaction of ceftolozane (pink) to protein encoded by PA4311.** Dashed yellow lines indicate hydrogen bonds.

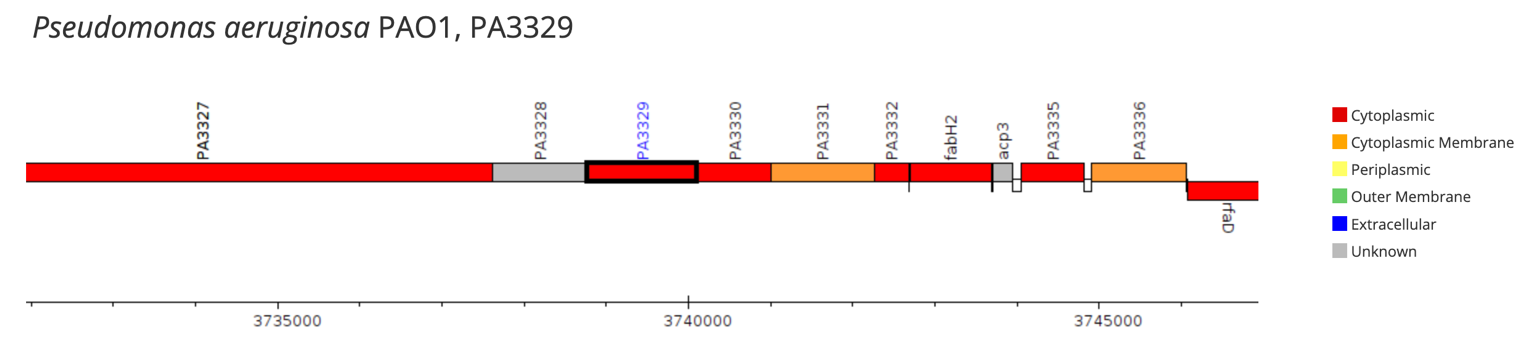

**Supplementary Figure 8.** **Genomic context of PA3329 and its neighbouring genes**. Screenshot obtained from <https://www.pseudomonas.com/feature/show?id=109475>^10^.

**Supplementary tables**

**Supplementary Table 1. Occurrence of common STs in C/T-susceptible and resistant groups.** STs are ordered by the total number of isolates in descending order.

| **ST** | **Susceptible (%)** | **Resistant (%)** |
| --- | --- | --- |
| 235 | 85 (52.1%) | 78 (47.9%) |
| 175 | 84 (77.1%) | 25 (22.9%) |
| 111 | 34 (47.2%) | 38 (52.8%) |
| 244 | 62 (89.9%) | 7 (10.1%) |
| 308 | 22 (37.3%) | 37 (62.7%) |
| 155 | 50 (89.3%) | 6 (10.7%) |
| 274 | 36 (83.7%) | 7 (16.3%) |
| 179 | 29 (76.3%) | 9 (23.7%) |
| 649 | 34 (94.4%) | 2 (5.6%) |
| 253 | 24 (77.4%) | 7 (22.6%) |
| 298 | 28 (100.0%) | 0 (0.0%) |
| 282 | 17 (70.8%) | 7 (29.2%) |
| 17 | 22 (95.7%) | 1 (4.3%) |
| 2211 | 22 (100.0%) | 0 (0.0%) |

**Supplementary Table 2. Carbapenemase and other beta-lactamase genes in C/T resistant and susceptible isolates.**

| **Genes** | **C/T resistant isolates (n=343)** | | **C/T susceptible isolates (n=1,339)** | |
| --- | --- | --- | --- | --- |
|  | **Count (%)** | **Genes** | **Count (%)** | **Genes** |
| *bla*_GES_ | 34 (9.9%) | *bla*_GES-5_ (18), *bla*_GES-1_ (11), *bla*_GES-20_ (4), *bla*_GES-19_ (3), *bla*_GES-26_ (1) | 4 (0.3%) | *bla*_GES-5_ (4) |
| *bla*_KPC_ | 4 (1.2%) | *bla*_KPC-2_ (3), *bla*_KPC-3_ (1) | 0 (0%) |  |
| *bla*_IMP_ | 79 (23.0%) | *bla*_IMP-4_ (31), *bla*_IMP-1_ (29), *bla*_IMP-14_ (8), *bla*_IMP_ (3), *bla*_IMP-8_ (2), *bla*_IMP-13_ (2), *bla*_IMP-7_ (2), *bla*_IMP-43_ (2), *bla*_IMP-75_ (1), *bla*_IMP-62_ (1) | 0 (0%) |  |
| *bla*_VIM_ | 49 (14.3%) | *bla*_VIM-2_ (25), *bla*_VIM-20_ (14), *bla*_VIM-1_ (6), *bla*_VIM-4_ (1), *bla*_VIM-47_ (1), *bla*_VIM-6_ (1), *bla*_VIM-5_ (1) | 0 (0%) |  |
| *bla*_NDM_ | 37 (69.1%) | *bla*_NDM-1_ (37) | 0 (0%) |  |
| *bla*_VEB_ | 8 (2.3%) | *bla*_VEB-9_ (4), *bla*_VEB-14_ (2), *bla*_VEB-5_ (1), *bla*_VEB_ (1) | 0 (0%) |  |
| *bla*_PER_ | 2 (0.6%) | *bla*_PER-1_ (2) | 0 (0%) |  |
| *bla*_OXA_ | 86 (25.1%) | *bla*_OXA-10_ (24), *bla*_OXA-2_ (21), bla_OXA_ (18), *bla*_OXA-14_ (8), *bla*_OXA-141_ (4), *bla*_OXA-210_ (3), *bla*_OXA-796_ (3), *bla*_OXA-1085_ (2), *bla*_OXA-4_ (2), *bla*_OXA-46_ (2), *bla*_OXA-15_ (1), *bla*_OXA-232_ (1), *bla*_OXA-101_ (1), *bla*_OXA-9_ (1), *bla*_OXA-415_ (1) | 91 (6.8%) | bla_OXA_ (57), *bla*_OXA-10_ (3), *bla*_OXA-2_ (22),  *bla*_OXA-46_ (1), *bla*_OXA-9_ (7), *bla*_OXA-677_ (1) |

**Supplementary Table 3. The occurrence of common mutations (present in** $\boldsymbol{\geq}$**5% of either C/T susceptible or resistant isolates) in previously described C/T resistance genes.** Mutations previously described as natural polymorphisms are indicated in plain text, mutations with known impact on C/T resistance are shown in bold.

| **Gene** | **C/T resistant isolates (n=115)** | | **C/T susceptible isolates (n=1332)** | |
| --- | --- | --- | --- | --- |
|  | **Count (%)** | **Common mutations (count)** | **Count**  **(%)** | **Common mutations (count)** |
| *ampC* | 109 (94.8%) | **V239A (9; 7.8%)** T105A (100; 87.0%)  G391A (34; 29.6%)  V205L (27; 23.5%)  G27D (22; 19.1%)  R79Q (19; 16.5%)  V356I (11; 9.6%)  L176R (10; 8.7%)  T21A (8; 7.0%)  A97V (8; 7.0%) | 1129 (84.8%) | **V239A (4; 0.3%)**  T105A (1101; 82.7%)  G391A (367; 27.6%)  V205L (347; 26.1%)  G27D (246; 18.5%)  R79Q (261; 19.6%)  V356I (101; 7.6%)  L176R (132; 9.9%)  T21A (106; 8.0%)  A97V (78; 5.9%) |
| *ampR* | 55 (47.8%) | **D135N (10; 8.7%)**  G283E (42; 36.5%)  M288R (40; 34.8%)  E114A (13; 11.3%) | 561 (42.1%) | **D135N (9; 0.7%)**  G283E (374; 28.1%)  M288R (362; 27.2%)  E114A (207; 15.5%) |
| *ampD* | 92 (80.0%) | G148A (80; 69.6%)  D183Y (30; 26.1%)  S175L (15; 13.0%)  R11L (12; 10.4%)  Q44H (6; 5.2%)  A136V (5; 4.3%) | 1051 (78.9%) | G148A (916; 68.8%)  D183Y (469; 35.2%)  S175L (72; 5.4%)  R11L (133; 10.0%)  Q44H (111; 8.3%)  A136V (90; 6.8%) |
| *mpl* | 91 (79.1%) | **M38fs (10; 8.7%)**  M297V (71; 61.7%)  A303V (13; 11.3%) | 949 (71.2%) | **M38fs (35; 2.6%)**  M297V (740; 55.6%)  A303V (217; 16.3%) |
| *dacB* | 39 (33.9%) | **G420S (8; 7.0%)**  A394P (18; 15.7%) | 339 (25.5%) | **G420S (0; 0.0%)**  A394P (212; 15.9%) |
| *mexR* | 56 (48.7%) | V126E (41; 35.7%) | 575 (43.2%) | V126E (466; 35.0%) |
| *nalC* | 107 (93.0%) | G71E (106; 92.2%)  S209R (74; 64.3%)  A186T (13; 11.3%)  E153Q (8; 7.0%)  A145V (8; 7.0%) | 1224 (91.9%) | G71E (1204; 90.4%)  S209R (866; 65.0%)  A186T (159; 11.9%)  E153Q (91; 6.8%)  A145V (124; 9.3%) |
| *nalD* | 17 (14.8%) |  | 210 (15.8%) |  |
| *ftsI* | 50 (43.5%) | **R504C (21; 18.3%)**  T91A (6; 5.2%) | 201 (15.1%) | **R504C (17; 1.3%)**  T91A (70; 5.3%) |
| *galU* | 3 (2.6%) |  | 28 (2.1%) |  |
| *oprD* | 77 (67.0%) | **frameshift indel**  **(26; 22.6%)**  **truncated (16; 13.9%)**  **premature stop E176* (8; 7.0%)** | 626 (47.0%) | **frameshift indel (249; 18.7%)**  **truncated (137; 10.3%)**  **premature stop E176* (1; 0.1%)** |

**Supplementary Table 4. GWAS-significant genes and their occurrence in machine learning models for C/T resistance prediction. Genes are ordered by decreasing occurrence in the Elastic Net model.**

| **Gene** | **Occurrence in**  **Elastic net** | ***P* value** | **Average**  **minimum allele frequency (MAF)** |
| --- | --- | --- | --- |
| *ftsI* | 10 | 5.05E-23 | 0.139808 |
| *ampR* | 10 | 3.52E-18 | 0.251644 |
| PA3329 | 9 | 1.47E-15 | 0.15295 |
| PA4311 | 7 | 6.86E-15 | 0.275563 |
| *rbdA* | 7 | 1.63E-10 | 0.01075 |
| *surE* | 7 | 9.24E-21 | 0.012 |
| *ampC* | 6 | 6.5E-15 | 0.354867 |
| PA3179 | 5 | 1.46E-10 | 0.139278 |
| PA4071 | 5 | 1.04E-12 | 0.204083 |
| *cdhA* | 5 | 3.11E-10 | 0.01 |
| *eno* | 5 | 7.35E-12 | 0.011 |
| PA1270 | 4 | 8.29E-10 | 0.065818 |
| *fusA1* | 3 | 1.43E-11 | 0.0376 |
| *braF* | 3 | 1.15E-10 | 0.059 |
| *coxA* | 3 | 1.74E-08 | 0.023 |
| PA2418 | 3 | 1.58E-13 | 0.325271 |
| PA4772 | 3 | 3.71E-09 | 0.119111 |
| PA2542 | 2 | 5.4E-12 | 0.4966 |
| PA4523 | 2 | 9.92E-09 | 0.01 |
| *gcvP2* | 2 | 2.91E-11 | 0.026 |
| PA1918 | 2 | 4.76E-11 | 0.01105 |
| *hsbR* | 1 | 5.85E-20 | 0.23299 |
| PA4736 | 1 | 5.31E-15 | 0.01345 |
| *hasS* | 1 | 6.55E-09 | 0.162 |
| PA1551 | 1 | 9.59E-12 | 0.0175 |
| *amaB* | 1 | 1.18E-11 | 0.01835 |
| PA0828 | 0 | 4.36E-15 | 0.031143 |
| *dgkA* | 0 | 8.34E-13 | 0.0265 |
| PA5524 | 0 | 4.12E-12 | 0.011 |
| *algF* | 0 | 3.71E-11 | 0.0111 |
| *rpsL* | 0 | 4.55E-11 | 0.0105 |
| *rpoS* | 0 | 6.34E-11 | 0.019 |
| PA0946 | 0 | 7.39E-11 | 0.0255 |
| PA3211 | 0 | 7.99E-11 | 0.023 |
| *accC* | 0 | 9.27E-11 | 0.016667 |
| PA0313 | 0 | 1.27E-10 | 0.1826 |
| *ppc* | 0 | 1.68E-10 | 0.01725 |
| PA0929 | 0 | 1.69E-10 | 0.0159 |
| *cntL* | 0 | 2.39E-10 | 0.034 |
| *cyaB* | 0 | 2.79E-10 | 0.0125 |
| *rpoC* | 0 | 4.64E-10 | 0.024 |
| PA4895 | 0 | 5.75E-10 | 0.108125 |
| PA2094 | 0 | 5.85E-10 | 0.106 |
| *algC* | 0 | 6.39E-10 | 0.025 |
| PA4929 | 0 | 6.65E-10 | 0.0124 |
| PA1232 | 0 | 6.96E-10 | 0.194111 |
| *pslG* | 0 | 7.34E-10 | 0.0187 |
| PA5504 | 0 | 1.18E-09 | 0.013 |
| PA4594 | 0 | 1.3E-09 | 0.3279 |
| PA1157 | 0 | 1.43E-09 | 0.031 |
| PA4520 | 0 | 1.57E-09 | 0.041 |
| *dnaA* | 0 | 2.11E-09 | 0.02 |
| PA4961 | 0 | 3.48E-09 | 0.019 |
| *pbpA* | 0 | 3.66E-09 | 0.035 |
| PA0345 | 0 | 3.89E-09 | 0.080667 |
| PA1923 | 0 | 3.99E-09 | 0.075 |
| PA4333 | 0 | 4.04E-09 | 0.026 |
| PA3287 | 0 | 4.43E-09 | 0.032 |
| *kinB* | 0 | 5.62E-09 | 0.0245 |
| *mucA* | 0 | 5.92E-09 | 0.02575 |
| *secD* | 0 | 8.86E-09 | 0.022 |
| PA0800 | 0 | 9.5E-09 | 0.017 |
| *tag* | 0 | 1.07E-08 | 0.0207 |
| PA4998 | 0 | 1.14E-08 | 0.03 |
| PA0032 | 0 | 1.24E-08 | 0.0117 |
| *thyA* | 0 | 1.24E-08 | 0.0117 |
| PA4338 | 0 | 1.25E-08 | 0.025 |
| *wapB* | 0 | 1.31E-08 | 0.03 |
| *amgS* | 0 | 1.6E-08 | 0.038 |
| PA1764 | 0 | 1.83E-08 | 0.0104 |
| PA3937 | 0 | 1.85E-08 | 0.0214 |
| *ampD* | 0 | 1.85E-08 | 0.113 |
| *phuR* | 0 | 2E-08 | 0.062 |

**Supplementary Table 5. Gene Ontology (GO) functions prediction for PA3329 and PA4311.**

| **Gene** | **Category** | **GO term** | **Predicted Probability** | **Annotation** |
| --- | --- | --- | --- | --- |
| PA3329 | Molecular function | GO:0003824 | 0.994 | catalytic activity |
|  | Cellular component | GO:0110165 | 0.979 | cellular anatomical structure |
|  |  | GO:0071944 | 0.891 | cell periphery |
|  | Biological process | GO:0008152 | 0.905 | metabolic process |
|  |  | GO:0009987 | 0.857 | cellular process |
| PA4311 | Molecular function | GO:0003824 | 0.964 | catalytic activity |
|  |  | GO:0016757 | 0.951 | glycosyltransferase activity |
|  |  | GO:0016758 | 0.929 | hexosyltransferase activity |
|  |  | GO:0016740 | 0.893 | transferase activity |
|  | Cellular component | GO:0110165 | 0.981 | cellular anatomical structure |
|  | Biological process | GO:0008152 | 0.913 | metabolic process |
|  |  | GO:0009987 | 0.902 | cellular process |
|  |  | GO:0009058 | 0.809 | biosynthetic process |
